# Association of caregiver nativity and U.S. residency on preschoolers’ time playing outdoors and screen time: Findings from the 2022 National Survey of Children’s Health

**DOI:** 10.64898/2026.05.07.26352664

**Authors:** Phoebe P. Tchoua, Sarah M. Peterson, Falon T. Smith, Tiwaloluwa A. Ajibewa, Emily Clarke, Erik A. Willis

**Author notes:** **Corresponding Author:** Phoebe P. Tchoua.

## Abstract

**Background:** Outdoor play and limited screen time are critical for preschoolers’ physical health and socio-emotional development, yet little is known about how caregiver nativity and acculturation shape these behaviors.

**Methods:** We analyzed the 2022-2023 National Survey of Children’s Health data for 10,157 U.S. children 3-5 years old. Generalized linear models estimated associations between caregiver nativity and length of U.S. residence and children’s outdoor play and weekday screen time, adjusting for child, caregiver, and household covariates. Models tested interactions with race/ethnicity.

**Results:** Overall, caregiver length of U.S. residence was not associated with children’s outdoor play. However, screen time differed – children whose caregivers arrived Pre-1997 had lower odds of screen time frequency, whereas those whose caregivers arrived between 1997-2005 had higher odds compared with children of U.S.-born caregivers. Associations for weekday outdoor play and screentime varied significantly by child race/ethnicity.

**Conclusions:** Caregiver length of U.S. residence appears more strongly related to preschoolers’ screen time than outdoor play, with notable differences across racial/ethnic groups. Culturally tailored strategies may be needed to reduce early childhood screen exposure and support healthy movement behaviors among immigrant families.

## INTRODUCTION

Physical activity through play is a fundamental component of a child’s development.^1^ In unstructured outdoor settings, it encourages movement at higher intensity levels (i.e., moderate to vigorous physical activity).^2^ The amount of time a child spends playing outdoors is directly related to increased health benefits (e.g., mental health, social-emotional well-being, prevention of myopia, etc.).^3^ Children should be active throughout the day and engage in a variety of movement behaviors, as recommended by the Physical Activity Guidelines for Americans.^4^ Experts further advise that preschool-aged children spend 60–90 minutes outdoors each day.^5^ However, recent national estimates indicate that most children play outdoors for one hour or less on weekdays and three hours or more on weekends.^6^ Time outdoors is also inversely related to other behaviors, such as screen time. More specifically, infrequent time outdoors is significantly associated with greater screen exposure,^7^ which, in turn, is linked to negative developmental and health-related outcomes, including increased adiposity.^8^ The American Academy of Pediatrics recommends limiting screen time for children aged 2-5 to a maximum of 1 hour per day,^9^ yet only 36% of children globally under six years of age meet this guideline,^9^ and more than 56% spend 2 hours per day on screens.^10^

It is well established that parents and caregivers play an essential role in shaping a child’s development and health by influencing their daily behaviors such as physical activity, outdoor play, and screen time. These behaviors are also influenced by broader ecological factors. Caregivers’ sociodemographic characteristics, including income, education, sex, and race/ethnicity, are all predictors of children’s time playing outdoors.^11–13^ For example, children tend to spend more time in outdoor play when their caregivers are physically active, value and engage in outdoor play, and understand its benefits.^11^ Higher levels of caregiver education and being part of a majority racial/ethnic group (e.g., non-Hispanic White) are also linked to increased outdoor play.^11^ On the other hand, caregiver characteristics linked to reduced outdoor play include being female, having overweight or obesity, having lower educational attainment, working full-time, belonging to a minority racial or ethnic group (e.g., African American/Black), and having recently immigrated.^12^ Social determinants of health, such as limited access to safe outdoor spaces, and economic constraints, help explain why many minority racial and ethnic groups experience reduced opportunities for outdoor play.^14–16^ In addition, higher levels of child screen time is associated with the race/ethnicity of the caregiver^7,13^ and increased sedentary time,^17^ but not the child’s age and sex.

Among these sociodemographic factors, caregiver nativity (i.e., U.S.-born or foreign-born) remains relatively understudied, yet may play an important role in shaping children’s physical activity and screen time behaviors. Of the existing studies, immigrants from high-income countries tend to report better physical health and health outcomes compared to native-born residents, even after controlling for age and education.^18,19^ Compared to children of native-born caregivers, children of foreign-born caregivers have more favorable health-related outcomes than would be expected based on their lower socioeconomic status (SES) or racial/ethnic minority background.^19^ However, this *healthy immigrant effect* advantage diminishes over time as immigrants’ length of stay in the new countries increases.^18^ For example, non-recent immigrants (i.e., migrated over ten years ago) in Canada have been shown to engage in fewer outdoor activities compared to those who immigrated more recently.^19^ These findings suggest that understanding the role of caregiver nativity in shaping children’s health behaviors warrants further exploration.

### Theoretical Framework

Over the years, researchers have used several theories and frameworks to understand influences on human health behavior. For example, the Bronfenbrenner Ecological Systems Theory (EST) posits that human development is shaped by nested and interrelated environmental systems at five levels, from proximal to distal.^20,21^ The most proximal level, the microsystem (e.g., family), directly influences the child; the mesosystem highlights the interaction between microsystems (e.g., community-family interaction); the exosystem refers to indirect influences through interactions with the microsystems and extended networks (e.g., local government, extended family network, mass media). At the macrosystem level, the influences are larger, non-tangible elements (e.g., cultural ideologies or expectations, social conditions, socioeconomic factors, acculturation), and the most distal system, the chronosystem, considers predictable and unpredictable changes over time in a child’s life (e.g., historical events, generational cultural changes).^20^

### The Current Study

While previous studies have looked at the influence of family, school, community, policy, and socioeconomic factors on children’s health behavior, less is known about acculturation and time in a new country. Specifically, research has yet to explore how the length of a caregiver’s residence in the U.S. may influence outdoor play and screen time in preschool-aged children. To date, no previous studies to our knowledge have investigated associations of time playing outdoors and in front of screens with the caregiver’s length of residence in the U.S. in preschool-aged children. While Dahl et al.^6^ reported the 2021 national level estimates of time spent outdoors and its association with various sociodemographic factors, acculturation measures (e.g., nativity, length of residence in the U.S.) were not included. This study aims to fill this gap by analyzing the 2022-2023 national-level data on time spent playing outdoors and in front of screens in children aged 3-5 years, looking at caregiver nativity and length of residence in the U.S. Grounded in Bronfenbrenner’s EST, we examined whether caregiver nativity and length of U.S. residence (nested within the macrosystem) were associated with outdoor play and screen time in preschool-aged children, and whether these associations varied by child’s sociocultural contexts in a nationally representative sample. We hypothesized that children of foreign-born caregivers (who immigrated recently) would exhibit healthier behaviors and that those whose caregivers had lived longer in the U.S. would show similar patterns to children of native-born caregivers.

## METHODS

### Study Design

This study is a secondary analysis of cross-sectional data from the 2022-2023 National Survey of Children’s Health (NSCH). According to https://childhealthdata.org “The Data Resource Center for Child and Adolescent Health, a project of the Child and Adolescent Health Measurement Initiative (CAHMI) supported by the Health Resources and Services Administration’s (HRSA) Maternal and Child Health Bureau (MCHB), provides online access to survey data from the NSCH. The DRC site allows users to compare state, regional, and nationwide results, and to access additional resources and personalized assistance for interpreting and reporting findings.” The survey provides comprehensive data on household caregiver demographics, children’s health and development, access to healthcare and support services, and neighborhood characteristics. For this study, we focused on caregiver characteristics and their responses regarding outdoor play behaviors among typically developing children aged 3-5 years. This study used publicly available secondary data from the NSCH; therefore, it was not considered human subjects research by our institutional review board.

### Study Participants

NSCH survey participants (i.e., one adult per household) first completed a screening questionnaire to determine household eligibility. Eligible households subsequently received one of three topical questionnaires based on the age of the randomly selected child: NSCH-T1 for children aged 0-5, NSCH-T2 for children aged 6-11, or NSCH-T3 for children aged 12-17. An adult caregiver familiar with the health status and needs of the randomly selected child completed the appropriate topical questionnaire. Households with children aged 0-5 were oversampled due to their typical underrepresentation in household surveys.^22^

Between July 2022 and January 2023, surveys were administered in English or Spanish in one of three formats: online, mailed paper survey, or telephone interview with telephone questionnaire assistance agents. In total, 67,269 (44.8% weighted completed rate) screening questionnaires with children ages 0 to 17 and 54,103 (30.9% weighted completed rate) detailed topical questionnaires were completed. Each record contains data on “the child’s health, special health care need status, health care, family functioning, parental health, neighborhood and community characteristics, health insurance coverage, and demographics.”.^23^

This study focused exclusively on children aged 3 to 5, as questions about outdoor play were only asked for this age group. Data were derived from the NSCH-T1, specifically from the following sections: “This Child’s Health Insurance Coverage (section E), “About You and This Child” (section H), “About Your Family and Household (Section I), “Child’s Caregivers” (Section J), and “Household Information” (Section K). The variables of interest included child and caregiver demographics, country of birth, length of stay in the U.S., time spent playing outdoors on weekdays and weekends, and time spent in front of a screen for children aged 3-5.

Children diagnosed with the following conditions were excluded from the analysis: Down syndrome, Cystic fibrosis, intellectual disability, Autism, and Cerebral Palsy (N=527). After excluding cases with missing data on caregiver place of birth and time in the U.S. (N=964), the final analytic sample consisted of 10,157 children.

### Measures

#### Dependent variables

Dependent variables consisted of: (1) time playing outdoors on weekdays (survey item: “ON MOST WEEKDAYS, how much time does this child spend playing outdoors?”), (2) time playing outdoors on weekends (survey item: ON AN AVERAGE WEEKEND DAY, how much time does this child spend playing outdoors?”), and (3) time in front of a screen (survey item: ON MOST WEEKDAYS, about how much time did this child spend in front of a TV, computer, cellphone or other electronic device watching programs, playing games, accessing the internet or using social media?”). Responses were recorded on a 5-point Likert scale: (1) less than 1 hour per day; (2) 1 hour per day; (3) 2 hours per day; (4) 3 hours per day; (5) 4 hours or more per day.

#### Independent variables

Independent variables included caregiver’s place of birth (born in the U.S., yes or no) and year caregiver arrived in the U.S. (recoded into four categories based on the caregiver’s year of arrival in the U.S.: Pre-1997, 1997-2005, 2006-2012, and 2013-2022).

#### Covariates

Variables included child age (3, 4, or 5 years old), sex (male or female), place of birth (inside or outside the U.S.), race and ethnicity (Hispanic, non-Hispanic [NH] White, NH Black, NH Asian, NH American Indian or Alaska Native, NH Native Hawaiian and Other Pacific Islander, or NH Multi-race), and insurance (public only [government assistance], private only [including ACA (Affordable Care Act) marketplace, employer, or TRICARE], or public and private). Hereafter, the term race will only be used, for example, White instead of NH White.

Variables related to the caregiver respondent who completed the questionnaire consisted of age (18-34, 35-54, 55-64, or 65+), sex (male or female), employment status (employed full-time, employed part-time, working without pay, not employed but looking for work, or not employed but not looking for work), and marital status (married, not married and living with a partner, never married, divorced, separated, or widowed). For the household highest level of education (less than high school, high school [including vocational, trade, or business school], some college or Associate degree, or college degree or higher), income (0-99% FPL, 100-199% FPL, 200-399% FPL, or 400% FPL or greater), and five neighborhood factors (presence of sidewalks or walking paths, presence of parks or playgrounds, absence of litter or garbage on the street or sidewalk, poorly kept or rundown housing, and vandalism such as broken windows or graffiti).

### Analysis

Descriptive analyses were conducted to summarize sample characteristics and estimate the weighted prevalence of children’s health behaviors. Generalized linear models (GLMs) using cumulative logistic regression were employed to examine associations between caregiver nativity (place of birth) and acculturation (years in the U.S.) and each outcome: weekday outdoor play, weekend outdoor play, and screen time. All models controlled for child’s sex, age, race, insurance type, household income, highest household adult education level, marital and employment status, and neighborhood factors. Interactions between race and the nativity/acculturation variable were tested, and significant interactions were further examined in race-stratified models (White, Black, Hispanic, Asian, and Multiracial; other groups were excluded due to small sample sizes). Reference groups included female sex, age 5 years, uninsured, ≥400% federal poverty level, college degree or higher education, and U.S.-born caregivers. Results are presented as unweighted frequencies (N), weighted percentages (%), adjusted odds ratios (AOR) with 95% confidence intervals (CIs), and p-values, with statistical significance defined as p<0.05. The models use cumulative logistic regression, where odds ratios greater than 1 indicate higher odds of being in a lower or equal category of the outcome. Thus, for outdoor time (where higher categories are healthier), AOR > 1 indicates less outdoor activity; for screen time (where higher categories are unhealthier), AOR > 1 indicates less screen use. All analyses were performed using SAS 9.4 (SAS Institute Inc., Cary, NC, USA).^24^

## RESULTS

### Descriptive Statistics

Most children ages 3-5 in the sample (N = 10,157) were born in the U.S. (97.79%, N = 9,958), identified as White (49.27%, N = 6,671), and had a U.S.-born caregiver (78.96%, N = 8,589). Full demographic characteristics of the child, the household, and caregiver respondent are presented in Table 1. The majority of children spent one hour or less outdoors on weekdays (35.43%, N = 3,134) and two hours outdoors on weekends (28.87%, N = 2,740). On most weekdays, one-third of children (33.77%, N = 3,316) spent two hours in front of a TV, computer, cellphone, or other electronic device for activities excluding schoolwork, such as watching programs, playing games, accessing the internet, or using social media.

**Table 1.** Characteristics of Children and Adults in the Sample (Weighted Estimates)

| Variable and Category | Frequency | Percent | SE of Percent |
| --- | --- | --- | --- |
| **Child** |  |  |  |
| **Age** |  |  |  |
| 3 | 3,490 | 33.69 | 0.97 |
| 4 | 3,301 | 33.56 | 0.94 |
| 5 | 3,366 | 32.75 | 0.92 |
| **Sex** |  |  |  |
| Male | 5,079 | 49.65 | 1 |
| Female | 5,078 | 50.35 | 1 |
| **Born in the U.S.** |  |  |  |
| Yes | 9,958 | 97.79 | 0.3 |
| No | 149 | 1.66 | 0.27 |
| No valid response | 50 | 0.6 | 0.14 |
| **Race** |  |  |  |
| Hispanic | 1,485 | 27.12 | 1.03 |
| White | 6,671 | 49.27 | 0.96 |
| Black | 492 | 10.50 | 0.68 |
| Asian | 615 | 4.88 | 0.35 |
| American Indian/Alaska Native | 52 | 0.54 | 0.1 |
| Native Hawaiian or Pacific Islander | 26 | 0.21 | 0.06 |
| Multi-race | 816 | 7.48 | 0.47 |
| **Time Outdoors: Weekdays** |  |  |  |
| 1 hour or less | 3,134 | 35.43 | 1.01 |
| 2 hours | 3,560 | 34.33 | 0.96 |
| 3 hours | 2,016 | 18.05 | 0.71 |
| 4 or more hours | 1,317 | 12.20 | 0.61 |
| **Time Outdoors: Weekend** |  |  |  |
| 1 hour or less | 1,785 | 21.94 | 0.93 |
| 2 hours | 2,740 | 28.87 | 0.92 |
| 3 hours | 2,596 | 24.18 | 0.82 |
| 4 or more hours | 2,905 | 25.01 | 0.81 |
| **Time Spent with**<br>**TV/Phone/Computer, most**<br>**weekdays** |  |  |  |
| Less than 1 hour | 1,824 | 16.59 | 0.72 |
| 1 hour | 2,884 | 26.57 | 0.82 |
| 2 hours | 3,316 | 33.77 | 1 |
| 3 hours | 1,321 | 14.19 | 0.7 |
| 4 or more hours | 741 | 8.88 | 0.61 |
| **Household**<br>**Health Insurance Type** |  |  |  |
| Public only | 2,190 | 30.34 | 1.02 |
| Private only | 7,120 | 58.48 | 1.03 |
| Private and public | 384 | 4.11 | 0.35 |
| Uninsured | 350 | 5.20 | 0.5 |
| **Household FPL** |  |  |  |
| 0–99% FPL | 1,131 | 17.88 | 0.91 |
| 100–199% FPL | 1,541 | 19.26 | 0.83 |
| 200–399% FPL | 2,973 | 28.35 | 0.88 |
| 400% FPL or greater | 4,512 | 34.51 | 0.86 |
| **Caregiver Respondent**<br>**Born in the U.S.** |  |  |  |
| Yes | 8,589 | 78.96 | 0.92 |
| No | 1,568 | 21.04 | 0.92 |
| **Arrival Cohort (if foreign-born)** |  |  |  |
| Pre-1997 | 397 | 22.29 | 1.95 |
| 1997–2005 | 373 | 27.15 | 2.66 |
| 2006–2012 | 401 | 24.46 | 2.06 |
| 2013–2022 | 397 | 26.10 | 2.08 |
| **Age** |  |  |  |
| 18–34 | 3,387 | 35.65 | 0.99 |
| 35–54 | 6,262 | 57.91 | 1.01 |
| 55–64 | 303 | 4.32 | 0.53 |
| 65+ | 172 | 2.12 | 0.29 |
| **Sex** |  |  |  |
| Male | 3,198 | 29.01 | 0.87 |
| Female | 6,932 | 70.64 | 0.87 |
| No valid response | 27 | 0.3 | 0.13 |
| **Marital Status** |  |  |  |
| Married | 8,209 | 75.52 | 0.96 |
| Not married, living with partner | 686 | 8.58 | 0.6 |
| Never married | 517 | 7.49 | 0.73 |
| Divorced | 411 | 3.82 | 0.35 |
| Separated | 166 | 2.49 | 0.34 |
| Widowed | 109 | 1.26 | 0.25 |
| No valid response | 59 | 0.8 | 0.17 |
| **Highest Education Among Adults** |  |  |  |
| Less than high school | 192 | 6.46 | 0.76 |
| High school or equivalent | 1,121 | 18.28 | 0.88 |
| Some college/associate degree | 1,962 | 18.73 | 0.77 |
| College degree or higher | 6,882 | 56.53 | 1.03 |
| **Employment Status** |  |  |  |
| Employed full-time | 6,750 | 62.60 | 0.96 |
| Employed part-time | 1,291 | 12.69 | 0.67 |
| Working without pay | 123 | 1.29 | 0.22 |
| Not employed, looking for work | 425 | 6.04 | 0.5 |
| Not employed, not looking | 1,503 | 16.45 | 0.7 |

### Model 1 – Associations between nativity, length of residence and weekday, weekend outdoor play, and screen time

On weekdays, children with foreign-born caregivers in the 1997–2005 cohort had higher odds of spending time outdoors compared to those with U.S.-born caregivers (AOR = 1.21, 95% CI = 0.84–1.74). In contrast, children in the 2006–2012 cohort showed slightly lower odds (AOR = 0.96, 95% CI = 0.69–1.33), while the cohorts arriving Pre-1997 and between 2013–2022 exhibited comparable odds to children with U.S.-born caregivers (see Table 2). None of these associations were statistically significant.

**Table 2.**
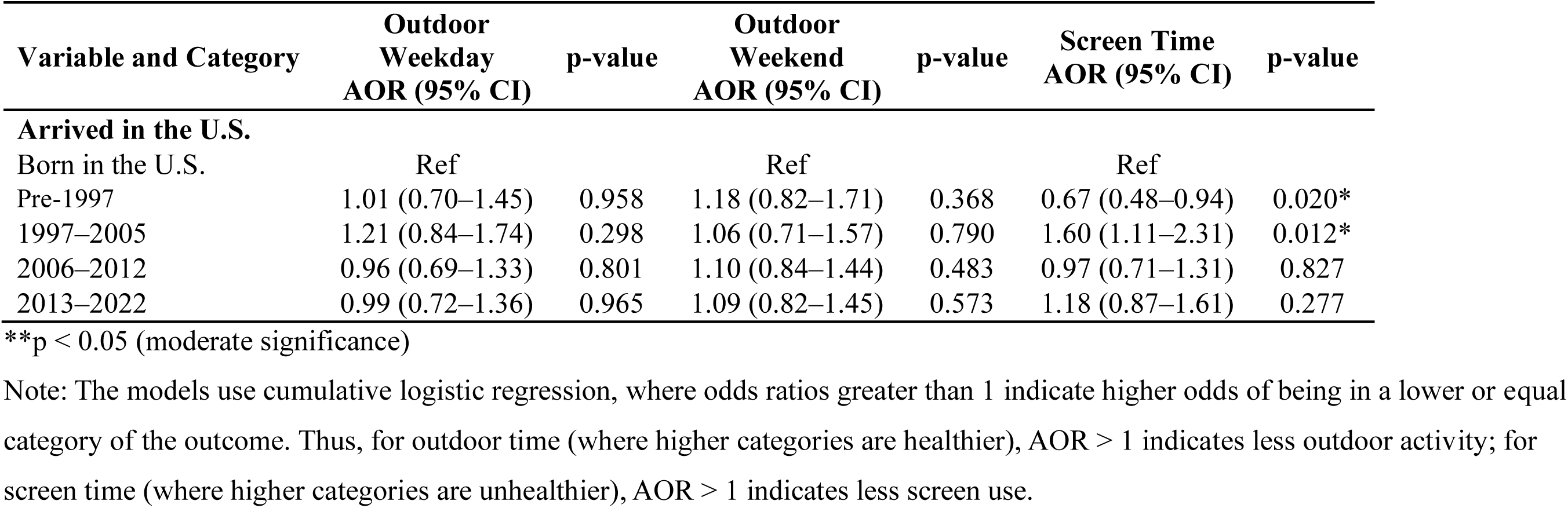
Adjusted Odds Ratios (AORs) and 95% Confidence Intervals for Outdoor Time and Screen Time by caregiver time in the U.S..

Overall, time spent outdoors during weekdays or weekends was not significantly associated with caregivers’ length of residence in the U.S (see Table 2).On weekends, children with foreign-born caregivers across all four arrival groups were similarly likely to spend less time outdoors than children with U.S.-born caregivers: Pre-1997 (AOR = 1.18, 95% CI = 0.82–1.71), 1997–2005 (AOR = 1.06, 95% CI = 0.71–1.57), 2006–2012 (AOR = 1.10, 95% CI = 0.84–1.44), and 2013–2022 (AOR = 1.09, 95% CI = 0.82–1.45) (see Table 2). None of these associations were statistically significant.

In contrast, screen time was significantly associated with caregivers’ length of residence in the U.S. Compared to children with U.S.-born caregivers, those with caregivers who arrived Pre-1997 spent significantly more time on screens (AOR = 0.67, 95% CI = 0.48–0.94), while those with caregivers who arrived between 1997 and 2005 were significantly less likely to do so (AOR = 1.60, 95% CI = 1.11–2.31) (see Table 2).

### Years in the U.S. by Race

Interaction analyses revealed that the association between length of residence in the U.S. and child behaviors differed significantly by race for time outdoor during weekdays (p = 0.007) and time in front of screens (p < 0.0001).

Time outdoor during the weekday and in front of screens differed significantly based on children’s race and their caregiver’s arrival to the U.S. compared to their peers with U.S. born caregivers. Among children with caregivers who migrated to the U.S. prior to 1997, White children had significantly higher odds of spending less time outdoors (AOR =2.17, 95% CI = 1.02–4.63), whereas multi-race children had significantly higher odds of spending time outdoors (AOR =0.53, 95% CI = 0.30–0.96). Screen time patterns in this cohort showed that White children (AOR =0.37, 95% CI = 0.14–0.96) and Asian children (AOR =0.32, 95% CI = 0.20–0.51) with foreign-born caregivers spent more time on screens. (See Table 3).

**Table 3.** Adjusted Odds Ratios (AORs) and 95% Confidence Intervals for Outdoor Weekday Activity and Screen Time, by Race and Years in the U.S.

| Race | Weekday AOR<br>(95% CI) | p-value | Screen Time<br>AOR (95% CI) | p-value |
| --- | --- | --- | --- | --- |
| Born in the U.S. | Ref | — | Ref | — |
| **White** |  |  |  |  |
| Pre-1997 | 2.17 (1.02–4.63) | 0.045* | 0.37 (0.14–0.96) | 0.041* |
| 1997–2005 | 0.65 (0.32–1.31) | 0.230 | 0.99 (0.56–1.73) | 0.961 |
| 2006–2012 | 0.60 (0.28–1.31) | 0.200 | 1.03 (0.45–2.34) | 0.95 |
| 2013–2022 | 1.30 (0.73–2.33) | 0.371 | 1.83 (0.98–3.40) | 0.058 |
| **Black** |  |  |  |  |
| Pre-1997 | 0.88 (0.34–2.27) | 0.792 | 0.54 (0.21–1.39) | 0.201 |
| 1997–2005 | 0.70 (0.28–1.76) | 0.451 | 6.93 (1.99–24.14) | 0.002** |
| 2006–2012 | 1.81 (0.68–4.83) | 0.237 | 0.32 (0.13–0.78) | 0.012** |
| 2013–2022 | 0.82 (0.35–1.92) | 0.642 | 1.18 (0.59–2.36) | 0.649 |
| **Hispanic** |  |  |  |  |
| Pre-1997 | 0.78 (0.46–1.31) | 0.347 | 1.01 (0.68–1.49) | 0.969 |
| 1997–2005 | 1.31 (0.80–2.15) | 0.290 | 1.18 (0.78–1.77) | 0.432 |
| 2006–2012 | 1.63 (0.96–2.77) | 0.068 | 0.88 (0.55–1.41) | 0.605 |
| 2013–2022 | 0.61 (0.37–0.99) | 0.045* | 1.83 (1.17–2.86) | 0.008** |
| **Asian** |  |  |  |  |
| Pre-1997 | 1.22 (0.67–2.22) | 0.524 | 0.32 (0.20–0.51) | <.001** |
| 1997–2005 | 2.38 (1.27–4.45) | 0.007** | 1.96 (1.19–3.21) | 0.008** |
| 2006–2012 | 0.52 (0.33–0.82) | 0.005** | 1.41 (0.90–2.22) | 0.138 |
| 2013–2022 | 1.20 (0.75–1.94) | 0.445 | 1.15 (0.70–1.91) | 0.574 |
| **Multi-race** |  |  |  |  |
| Pre-1997 | 0.53 (0.30–0.96) | 0.035* | 0.67 (0.35–1.29) | 0.231 |
| 1997–2005 | 0.70 (0.36–1.39) | 0.310 | 4.84 (1.81–12.98) | 0.002** |
| 2006–2012 | 1.67 (0.57–4.93) | 0.349 | 2.16 (0.82–5.74) | 0.120 |
| 2013–2022 | 2.54 (1.38–4.69) | 0.003** | 0.19 (0.11–0.35) | <.001** |

\*p < 0.01 (strong significance)
\*\*p < 0.05 (moderate significance)
Note: The models use cumulative logistic regression, where odds ratios greater than 1 indicate higher odds of being in a lower or equal category of the outcome. Thus, for outdoor time (where higher categories are healthier), AOR > 1 indicates less outdoor activity; for screen time (where higher categories are unhealthier), AOR > 1 indicates less screen use.

In the 1997-2005 cohort, Asian children had significantly higher odds of spending less time outdoors (AOR =2.38, 95% CI = 1.27–4.45). Black (AOR =6.93, 95% CI = 1.99–24.14), Asian (AOR =1.96, 95% CI = 1.19–3.21), and multi-race (AOR =4.84, 95% CI = 1.81–12.98) children were significantly more likely to spent less time on screens. In the 2006-2012 cohort, Asian children spent significantly more time outdoor (AOR =0.52, 95% CI = 0.33–0.82), and Black children reported spending significantly more time on screens (AOR =0.32, 95% CI = 0.13–0.78). (See Table 3).

In the 2013-2022 cohort, Hispanic children spent significantly more time outdoors (AOR =0.61, 95% CI = 0.37–0.99), whereas multi-race children spent significantly less time outdoor (AOR =2.54, 95% CI = 1.38–4.69). Screen time patterns were mixed, Hispanic children spent significantly less time on screens (AOR =1.83, 95% CI = 1.17–2.86), and multi-race children spent significantly more time on screens (AOR =0.19, 95% CI = 0.11–0.35). (See Table 3).

## DISCUSSION

This is the first study to our knowledge that examined caregiver nativity and length of U.S. residence in relation to preschool-aged children’s outdoor play and screen time using nationally representative data. Overall, caregiver length of residence was not significantly associated with children’s time playing outdoors during weekdays or weekends. However, screen time varied, with children whose caregivers arrived in the U.S. between 1997-2005 spending more time on screens compared to older cohorts who arrived Pre-1997. Additionally, the effect of length of residence in the U.S. on the likelihood of time playing outdoors on weekdays and screen time varied significantly by race. Interpreted through Bronfenbrenner’s EST. These findings suggest that acculturation (operationalized as length of residence) reflects macrosytem-level cultural adaptation processes that may influence sedentary behaviors more strongly than outdoor play by influencing norms, expectations, and routines within children’s microsystems. Race further interacts with these macrosystem factors, suggesting that both acculturation and racial/ethnic contexts may contribute to the differences observed in health behavior of children with native- and foreign-born caregivers.

### Time playing outdoors

Caregiver length of U.S. residence was not significantly associated with children’s outdoor play on either weekdays or weekends. Previous studies have found that the presence or absence of sidewalks, green space, neighborhood safety, and geographical location impacted time outdoors.^11,25^ Children living in metropolitan statistical areas had a lower prevalence of time outdoors.^6,11^ From an ecological perspective, these factors reflect exosystem conditions (e.g., sidewalks, green space) and macrosystem influences(e.g., cultural norms about outdoor play, social conditions, socioeconomic factors) that may exert stronger influence on outdoor play than caregiver’s acculturation alone. The absence of an association between caregiver years in the U.S. and outdoor play in this study may also be due to sample size; a larger and more diverse sample of foreign-born caregivers may reveal different patterns. Additionally, several key exosystem and macrosystem influences on outdoor play (e.g., access to green space, geographical location, work schedules, weather) were not measured and should be incorporated in future studies.

We found that, compared to children of U.S.-born caregivers, children in both the pre-1997 and 2013–2022 cohorts spent similar amounts of time playing outdoors. In contrast, the 1997–2005 cohort spent less time outdoors, whereas the 2006–2012 cohort spent more. Interpreted through Bronfenbrenner’s EST, these cohort differences may reflect shifts in macrosystem-level cultural norms and exosystem-level environmental conditions that shaped immigrant families’ engagement with outdoor spaces over time. Unlike the findings of Conrad et al^26^, who reported that immigrant children in Germany spent more time indoors and less time outdoors than non-immigrant children. Our results suggests that macrosystem and exosystem factors, such as cultural norms, immigration climate, neighborhood structures, safety perceptions, and economic conditions, likely varied across arrival cohorts, adding complexity to the interpretation of these patterns. It is important to note that outdoor play is influenced more by environmental factors than cultural adaptation alone. Given the limited research on outdoor play, nativity, and immigration, additional studies are needed in pre-school aged children, with consistent measurement of outdoor play and age-specific analyses rather than broad grouping (e.g., 0-12 vs. 0-2, 3-5, 6-10, 11-12).

### Screen time

In our sample, both caregiver duration of U.S. residence and the child’s race were significantly associated with children’s screen time. Children in the oldest cohort, pre-1997, had the most amount of screen time, whereas those from the 1997-2005 cohort had the least screen time, compared to children with native-born caregivers. This differs from our hypothesis that children of foreign-born with the longest duration in the U.S. would exhibit screen time behaviors similar to those of native-born caregivers. Interpreted through Bronfenbrenner’s EST, these cohort differences likely reflect macrosystem-level shifts in cultural norms and technological environments, as well as the exosystem-level changes in the availability and accessibility of digital media. The rapid expansion of digital technology (e.g., Wi-Fi, smartphone, tablets, streaming services, etc.) over the past three decades has altered the broader ecological context in which families make decision about screen use, and this may partially explain cohort differences in screen exposure. Because caregiver behavior strongly influences that of the child, perhaps caregivers who immigrated between 1997 to 2005 may have developed screen-use patterns in a macrosystem with fewer digital demands and access and may therefore model lower screen use for their children. Zhang et al.^27^ found that Latino fathers who made positive behavioral changes, such as spending less time on screens, positively influenced their children’s screen time behaviors, and other studies show that greater parental screen use increased the likelihood of their children exceeding the daily screen time limit.^11^

### Race/Ethnicity findings

In our study, variation by race and caregiver’s time in the U.S. was observed; however, no consistent linear pattern emerged between increasing caregiver’s length of residence in the U.S. and children’s outdoor play. White children in the pre-1997 cohort spent less time outdoors, whereas Asian children showed mixed patterns, with the 1997-2005 cohort spending less time outdoors and the 2006-2012 cohort spending more time outdoors. Multi-race children also demonstrated divergent patterns, with the Pre-1997 cohort spending more time outdoors but less time in the 2013-2022 cohort. Hispanic children in the 2013-2022 cohort spent more time outdoors. These heterogeneous patterns suggest that the relationship between nativity, race, and outdoor play is not uniform across groups or historical periods.

Interpreted through Bronfenbrenner’s EST, these findings suggest that racial/ethnic and cohort differences in outdoor play reflect an interaction between macrosystem-level cultural norms (e.g., beliefs about outdoor play, acculturation) and exosystem-level environmental conditions (e.g., neighborhood safety, sidewalk availability, access to green space) that differ across racial groups and immigration cohorts. Gavryutina et al.^28^ found that caregivers from minority racial and ethnic groups were less likely to view outdoor play as an important cultural value and reported fewer community members engaging in outdoor activities due to cultural norms and family traditions, highlighting the role of macrosystem influences. At the same time, structural barriers such as neighborhood safety and access to green space, both exosystem factors, may limit outdoor play opportunities for certain populations. Together, these studies support the ecological interpretation that children’s outdoor play is shaped by the combined influence of cultural values, structural environments, and historical context rather than by caregiver acculturation alone, underscoring the importance of culturally relevant messaging about time outdoors so families from diverse racial and ethnic backgrounds can embrace its benefits.

Screen time patterns varied substantially by race/ethnicity and immigration cohort. Compared to children of U.S.-born caregivers, screen time use for White and Asian children in the pre-1997 cohort was higher, and lower for Black, Asian, and multi-race children in the 1997–2005 cohort. Black children in the 2006–2012 cohort had higher screen use, while in the 2013–2022 cohort, Hispanic children had lower screen use and multiracial children had higher use. A systematic review by Veldman et al.^13^ found that the relationship between screen time and variables such as age and ethnicity among children aged 0-5 years varied across studies, with some reporting an association and others reporting none. Taken together, these mixed patterns suggest that screen time behaviors are shaped by a combination of cultural norms, family routines, and structural conditions rather than caregiver acculturation alone.

## Strengths and limitations

Our study has several strengths and limitations. To our knowledge, this is the first study to examine caregiver nativity and length of U.S. residence in relation to preschool-aged children’s outdoor play and screen time. Our analyses highlight that the factors influencing children’s outdoor play behaviors are complex and do not always follow a linear or predictable pattern. The study is in part limited due to its cross-sectional nature and relies on self-reported data, which are subject to responder biases (e.g., social desirability) and recall challenges. Additionally, the NSCH does not capture key acculturation-related variables, such as caregiver language proficiency, region of origin, or voluntary versus forced migration, that may shape caregiver behaviors and children’s activity patterns. As a result, this study operationalized caregiver acculturation using length of U.S. residence. While this metric is commonly employed in research, it may not fully capture individuals’ perspectives on integration, assimilation, separation, or marginalization.

## Conclusion

The results of this study revealed that caregiver length of residence in the U.S. was not significantly associated with children’s outdoor play on either weekdays or weekends. In terms of screen time, children whose caregivers arrived in the U.S. between 1997-2005 spent more time on screens compared to those whose caregivers arrived Pre-1997. The influence of caregiver length of residence on children’s weekday outdoor play and screen time differed significantly by race. These findings suggest that both caregiver nativity-related factors and child race should be considered when designing future research and developing health promotion programs aimed at improving outdoor play and reducing screen time among young children.

## NEW CONTRIBUTION TO THE LITERATURE

Until now, no studies have investigated the relationship between preschool aged children time spent playing outdoors or in front of screens and their caregiver’s length of residence in the U.S. This study addresses that gap by exploring how caregiver nativity and duration of U.S. residence are associated with children’s outdoor play and screen time, and how these associations differ across sociocultural contexts within a nationally representative sample. The findings underscore the need for further research focused on this population, which can inform public health professionals in developing more culturally relevant health promotion and disease prevention strategies.

## Acknowledgments

This research was conducted with support and resources provided by the Odum Institute for Research in Social Science at UNC-Chapel Hill.

## Statements and Declarations Funding

First author, Dr. Tchoua, was supported by the T32 Cancer Health Disparities Training Grant (T32CA128582) from the National Cancer Institute of the National Institutes of Health. The authors declare they have no financial interests.

## Competing Interests

Phoebe P. Tchoua, Sarah M. Peterson, Falon T. Smith, Tiwaloluwa A. Ajibewa, Emily Clarke, and Erik A. Willis declare they have no financial interests.

## Author Contribution

Conceptualization: Phoebe P. Tchoua; Methodology: Phoebe P. Tchoua; Formal analysis and investigation: Phoebe P. Tchoua; Writing - original draft preparation: Phoebe P. Tchoua; Writing - review and editing: Phoebe P. Tchoua, Sarah M. Peterson, Falon T. Smith, Tiwaloluwa A. Ajibewa, Emily Clarke, Erik A. Willis; Supervision: Erik A. Willis

## Ethics Approval

This study used publicly available secondary data from the NSCH; therefore, it was not considered human subjects research by our institutional review board.

## Data Availability

The data that support the findings of this study are available at https://www.childhealthdata.org/help/dataset

## Notes

### Competing Interest Statement

The authors have declared no competing interest.

### Funding Statement

First author was supported by the T32 Cancer Health Disparities Training Grant (T32CA128582) from the National Cancer Institute of the National Institutes of Health. The authors declare they have no financial interests.

### Author Declarations

The study used ONLY openly available human data that were originally located at: https://www.childhealthdata.org/help/dataset

